# The necessity of pyloric drainage in esophagectomies: protocol of a meta-analysis and a systematic review of randomized controlled trials

**DOI:** 10.1101/2022.08.24.22279164

**Authors:** Armand Csontos, Dávid Németh, Lajos Szakó, Gergő Berke, Dóra Lili Sindler, Péter Hegyi, András Papp

**Affiliations:** Institute for Translational Medicine, University of Pécs, Medical School, Pécs, Hungary, H-7624 Pécs, Szigeti út 12; Department of Surgery, University of Pécs, Medical School, Pécs, Hungary, H-7624 Pécs, Ifjúság útja 13; Institute of Bioanalysis, University of Pécs, Medical School, Pécs, Hungary, H-7624 Pécs, Honvéd utca 1; János Szentágothai Research Centre, University of Pécs, Medical School, Pécs, Hungary, H-7624 Pécs, Ifjúság útja 20; Department of Emergency Medicine, University of Pécs, Medical School, Pécs, Hungary, H-7624 Pécs, Ifjúság útja 13; First Department of Medicine, University of Szeged, Medical School, Szeged, Hungary, H-6725, Szeged Korányi fasor 8-10; Hungary Centre for Translational Medicine, Semmelweis University, Budapest, Hungary, H-1085 Budapest, Üllői út 26

**Keywords:** pyloroplasty, pyloromyotomy, oesophagectomy, eosophageal cancer

## Abstract

**Background:** Esophageal carcinoma is the 8th most common malignant tumour in the world with more than 600 000 cases (3.1% of all), while being the 6th most common reason of tumour mortality, causing more than 500 000 deaths (5.5% of all) annually. The 1, 3 and 5 year-prevalence are 2.4%, 1.6% and 1.3% respectively. The question of this meta-analysis is whether pyloric drainage is preferable over the lack of pyloric drainage during elective esophagectomies in patients suffering from esophageal cancer, regarding mortality, anastomosis leakage, respiratory morbidity, vomiting, gastric emptying time.

**Methods:** We plan to identify randomized controlled trials to investigate the question by performing extensive search in multiple databases. Based on of predefined criteria, two independent authors will perform the steps of selection, after which appropriate statistical analysis will be performed to identify potential significant differences. Cochrane Risk of Bias Tool 2, and GRADE approach will be used to estimate the risk of bias and quality of results.

**Dissemination plans:** We plan to distribute our results in peer-reviewed journal.

## Introduction

Esophageal carcinoma is the 8th most common malignant tumour in the world with more than 600 000 cases (3.1% of all) and it is the 6th most common reason of tumour mortality, causing more than 500 000 deaths (5.5% of all) annually. The 1, 3 and 5 year-prevalence are 2.4%, 1.6% and 1.3% respectively [1].

The 5-year survival is still low in patients with tumours, which is 19% in the USA [2] and 12% in Europe [3]. The outcome of the surgical treatment depends on the stage of the tumour, the patient’s condition, and the skill of the surgeon [4], therefore the outcomes may vary.

The 8th edition of UICC-AJCC TNM Classification suggests to treat esophageal cancer in the stage of I-IIB by esophagectomy [5]. For a long time, intraoperative pyloric drainages were routine procedures during elective esophagectomies in esophageal carcinoma, to protect the patients from postoperative complications of anastomosis insufficiency, aspiration, gastric emptying. There are some articles in the literature investigating the effects of pyloric drainage procedure, but there is no clear conclusion about the usefulness of intraoperative pyloroplasty and some limitations arise [6-8].

The question of this meta-analysis is whether pyloric drainage is preferable compared to the lack of pyloric drainage during elective esophagectomies in patients suffering from esophageal cancer, investigating mortality, anastomosis leakage, respiratory morbidity, vomiting, gastric emptying time.

## Methods

### Participants/population

Patients, who underwent intraoperative esophagectomy due to esophageal cancer are eligible for inclusion. We excluded those patients, on whom esophagial resection was performed due to any other cause.

### Interventions, exposures

The interventions of this analysis are the different types of intraoperative pyloric drainage, including pyloromyotomy and pyloroplasty.

### Comparators/control

The control in the case of this analysis is the lack of any kind of pyloric drainage.

### Main outcomes

We plan to investigate the mortality, anastomosis leakage, respiratory morbidity, vomiting, gastric emptying time.

### Search strategy

We will search the following databases: MEDLINE (via PubMed), Embase, Cochrane Library, Web of Science, and Scopus, with the following search key: “(((upper GI OR upper gastrointestinal OR esophagus OR oesophagus OR esophageal OR oesophageal OR stomach OR gastric) AND (surgery OR surgical OR operative OR operation OR resection)) OR (esophagectomy OR oesophagectomy OR gastrectomy)) AND drain*”. We do not intend to use any restrictions. Only randomized controlled trials will be included. Any other type of publication will be excluded from the analysis.

### Data extraction

Two independent reviewers (A.C., L.S.) will perform the selection first by title, second by abstract, last by full text following pre-discussed aspects. Data extraction will be done by the same two independent reviewers onto a pre-established Excel (Office 365, Microsoft, Redmond, WA, USA) worksheet. Extracted data consists of year of publication, name of the first author, study design, applied surgical modalities, demographic data, mortality, anastomosis leakage, respiratory morbidity, vomiting and gastric emptying time. Disagreements regarding both selection and data extraction will be resolved by consensus.

### Strategy for data synthesis

We plan to use the Comprehensive Meta-Analysis (Version 3) software (Biostat, Inc., Engelwood, MJ, USA) for meta-analytic calculations. During the data synthesis the working group of the Cochrane Collaborations recommendations will be used. We will calculate pooled odds ratios (ORs) with their 95% confidence intervals (CIs) from raw data in the case of dichotomous variables. In the case of continuous variables, weighted mean differences (WMD) will be calculated with their 95% confidence intervals. The random effect model with the estimation of DerSimonian and Laird [9] will be used. To assess heterogeneity, Cochrane’s Q and the I2 statistics will be used. Statistical significance will be declared in the case of P < 0.05. We will examine the publication bias By visual inspection of funnel plots. We plan to perform a trial sequential analysis to assess the necessary number of cases to obtain conclusive evidence in each outcome using the trial sequential analysis tool from Copenhagen Trial Unit (Centre for Clinical Intervention Research, Denmark).

### Quality assessment

To assess the risk of bias and quality of results we will use the Cochrane Risk of Bias Tool 2, and GRADE approach respectively.

The work process to be performed is presented in the following schematic diagram (Figure1).

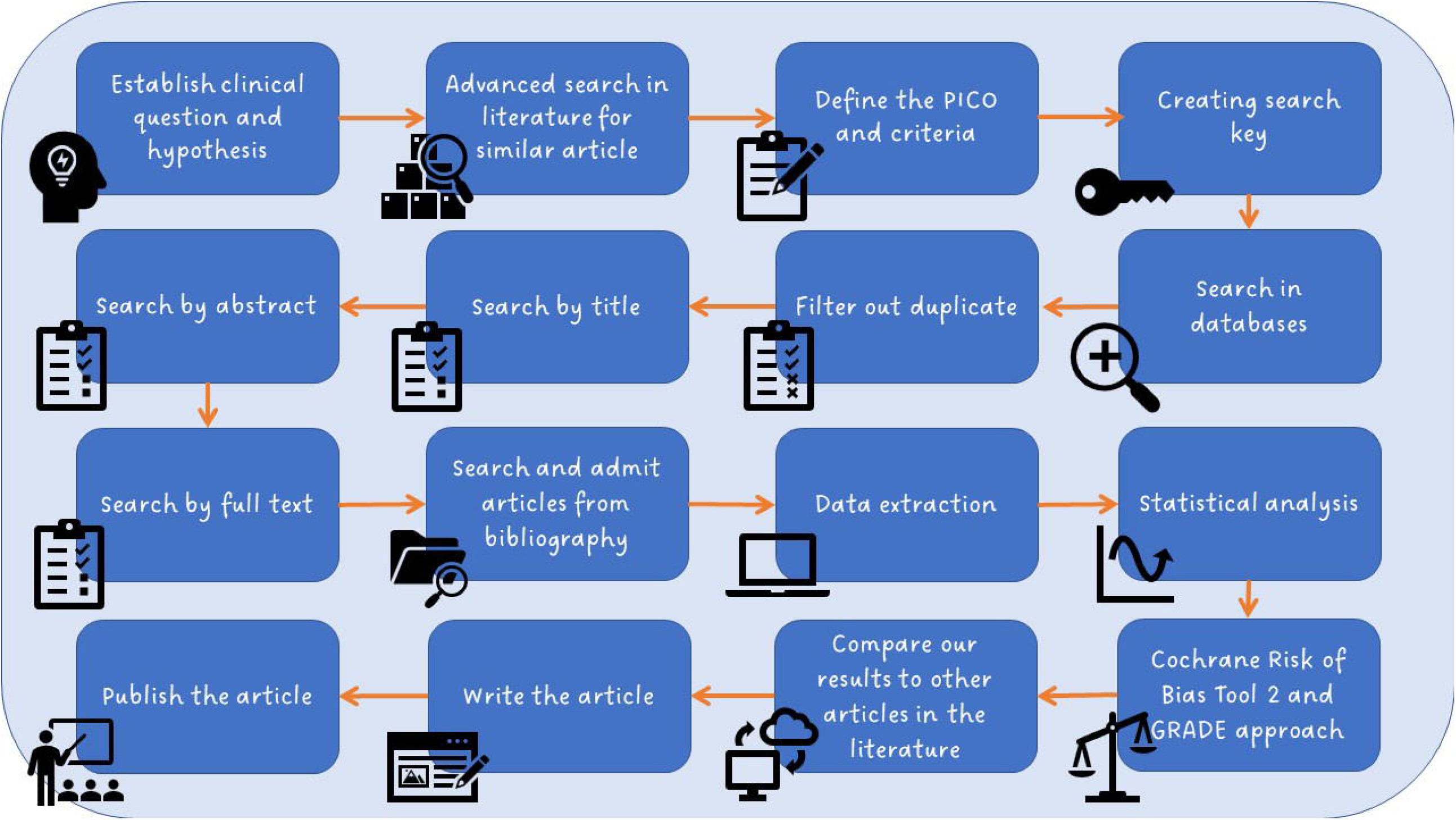

## Discussion

During elective esophageal surgeries in patients with esophageal carcinoma, pyloroplasty following esophagectomy was a routine procedure for a long time [10, 11], but nowadays minimal invasive procedures become increasingly available, therefore, and based on previous works intraoperative pyloroplasty may not be associated with benefits.

Pyloroplasty itself is an additional intervention, thus it can prolong the operation time, and it can be associated with general perioperative complications, therefore the question of the necessity of the pyloroplasty arises again.

Minimal invasive esophagectomy (MIE) has many advantages, therefore is becoming more widespread. Comparison of laparoscopic, thoracoscopic, totally minimally invasive and robotic esophagectomy shows decreased perioperative morbidity and hospitalization time against the open surgery. MIE is not detrimental even to perioperative mortality [12].

If the symptom of gastric stasis occurs, balloon dilatation, pyloric bouginage, endoscopic myotomy, botulinum toxin injection, erythromycin medication or per-oral gastric pyloromyotomy (GPOP) [10, 13-16] can be performed. All these new postoperative methods are safe and accessible procedures become more popular.

The topic is becoming more popular in the literature recently. Between the pyloroplasty and the control groups, Arya et al. [6], Gaur et al. [8] and Khan et al. [11] could not show significant difference in most outcomes in their work, however this research had some limitations, due to the small number of patients and heterogeneity of the definitions of outcomes in the enrolled studies.

We plan to investigate the question again, with rigorous inclusion and selection criteria enrolling only randomized controlled trials, using multiple data bases with the most specific search key.

We suspect that clear significant differences will be shown by using appropriate statistical analysis. Cochrane Risk of Bias Tool 2, and GRADE approach we will perform to estimate the risk of bias and quality of results.

Our dissemination plan is to publish our results to peer-reviewed high-quality journals.

## Data Availability

All data produced in the present study are available upon reasonable request to the authors

